# Acute pharmacodynamic responses to sitagliptin: Drug-induced increase in early insulin secretion in oral glucose tolerance test

**DOI:** 10.1101/2023.09.24.23296026

**Authors:** Amber L. Beitelshees, Elizabeth A. Streeten, Zhinous Shahidzadeh Yazdi, Hilary B. Whitlatch, Braxton D. Mitchell, Alan R. Shuldiner, May E. Montasser, Simeon I. Taylor

**Affiliations:** Department of Medicine, Division of Endocrinology, Diabetes, and Nutrition, University of Maryland School of Medicine, Baltimore, MD 21201, USA

## Abstract

**Aim:** DPP4 inhibitors are widely prescribed as treatments for type 2 diabetes. Because drug responses vary among individuals, we initiated investigations to identify genetic variants associated with the magnitude of drug responses.

**Methods:** Sitagliptin (100 mg) was administered to 47 healthy volunteers. Several endpoints were measured to assess clinically relevant responses – including the effect of sitagliptin on glucose and insulin levels during an oral glucose tolerance test (OGTT).

**Results:** This pilot study confirmed that sitagliptin (100 mg) decreased the area under the curve for glucose during an OGTT (p=0.0003). Furthermore, sitagliptin promoted insulin secretion during the early portion of the OGTT as reflected by an increase in the ratio of plasma insulin at 30 min divided by plasma insulin at 60 min (T30:T60) from 0.87+/-0.05 to 1.62+/-0.36 mU/L (p=0.04). The magnitude of sitagliptin’s effect on insulin secretion (as judged by the increase in the T30:T60 ratio for insulin) was correlated with the magnitude of sitagliptin-induced increase in the area under the curve for intact plasma GLP1 levels during the first hour of the OGTT. This study confirmed previously reported sex differences in glucose and insulin levels during an OGTT. Specifically, females exhibited higher levels of glucose and insulin at the 90-180 min time points. However, we did not detect significant sex-associated differences in the magnitude of sitagliptin-induced changes in T30:T60 ratios for either glucose or insulin.

**Conclusions:** T30:T60 ratios for insulin and glucose during an OGTT provide useful indices to assess pharmacodynamic responses to DPP4 inhibitors.

## 1 | INTRODUCTION

The past two decades have been a golden age for discovery and development of diabetes drugs – including three classes of innovative drugs: dipeptidylpeptidase-4 (DPP4) inhibitors, GLP1 receptor agonists, and sodium-glucose cotransporter-2 (SGLT2) inhibitors ^1^. There are at least four approved DPP4 inhibitors: sitagliptin, saxagliptin, vildagliptin, and linagliptin ^1^.

Sitagliptin was the first drug in the class to be approved and remains the most widely prescribed dipeptidylpeptidase-4 (DPP4) inhibitor. According to FDA-approved prescribing information, sitagliptin decreases mean HbA1c by 0.7% when administered to type 2 diabetic patients who are inadequately controlled with metformin. However, there is substantial inter-individual variation in the magnitude of HbA1c-lowering. While some patients experience very modest HbA1c-lowering, others experience HbA1c-lowering exceeding 1.5% ^2^.

DPP4 inhibitors are not the most efficacious class of diabetes drugs with respect to HbA1c-lowering ^1,3–5^, but they have the most favorable profiles with respect to safety and tolerability. Unlike GLP1 receptor agonists ^1^, they do not commonly cause nausea or vomiting. Unlike SGLT2 inhibitors ^6^, they are not associated with an increased risks of genital infections or ketoacidosis. Alogliptin is already available as a generic drug. When other DPP4 inhibitors experience loss of marketing exclusivity in the relatively near future, DPP4 inhibitors will represent an affordable oral antidiabetic drug that can be prescribed early in the course of type 2 diabetes to patients who are not adequately controlled on metformin. In this context, it would be desirable to be able to predict which patients are likely to experience above-average glycemic responses to DPP4 inhibitors. For example, if a patient experiences above-average HbA1c-lowering (>0.7%) in response to a DPP4 inhibitor, that degree of glycemic improvement would provide substantial clinical benefit to the patient. According to GoodRx, a one-month supply of sitagliptin (100 mg) costs ∼$550 in the US whereas a one-month supply of generic alogliptin (25 mg) is available for ∼$150 (i.e., ∼70% lower price). The price of generic DPP4 inhibitors is projected to fall to levels comparable to generic sulfonylureas or metformin as a result of competition after sitagliptin and other DPP4 inhibitors experience loss of marketing exclusivity. In light of the high degree of safety and tolerability, generic DPP4 inhibitors have potential to displace generic sulfonylureas and generic pioglitazone in many patients’ therapeutic regimens – especially if it becomes possible to identify individual patients likely to experience above-average HbA1c-lowering.

This manuscript describes an approach to assess the magnitude of an individual’s acute pharmacodynamic response to sitagliptin by analyzing data from an oral glucose tolerance test (OGTT). Whereas sitagliptin does not exert a significant effect on the area under the curve (AUC) for glucose-stimulated insulin secretion, sitagliptin administration shifts the time course of insulin secretion to earlier times. Similarly, sitagliptin significantly alters the time course for glucose levels during an OGTT. These data provide validation for a promising approach to conduct a pharmacogenomic study to identify genetic variants that are associated with pharmacodynamic responses to sitagliptin.

## 2 | METHODS

### 2.1 | Study population: recruitment and screening

The Old Order Amish population of Lancaster County, PA emigrated from Central Europe in the early 1700’s. University of Maryland School of Medicine researchers have been studying genetic determinants of cardiometabolic health in this population since 1993. To date, ∼10,000 Amish adults participated in one or more studies as part of the University of Maryland Amish Research Program (http://www.medschool.umaryland.edu/endocrinology/Amish-Research-Program/). The present study was a sub-study of a pilot study in healthy volunteers in which we investigated pharmacodynamic responses to exenatide (NCT05071898 at clinicaltrials.gov) ^7^. All procedures were reviewed and approved by the University of Maryland Baltimore Institutional Review Board.

A research nurse, accompanied by a liaison (a member of the Amish community), made home visits to invite individuals to participate in the study. If individuals expressed interest, the study was explained in detail; potential participants were invited to sign an informed consent form.

Thereafter, the research nurse obtained a medical history; measured height, weight, and blood pressure; and obtained blood samples for screening laboratory tests (hematocrit, fasting plasma glucose, serum creatinine, serum AST, serum ALT, serum TSH, and HbA1c).

### 2.2 | Study design: overview

Participants in the parent clinical trial with exenatide (NCT05762744) were offered the opportunity to participate in this clinical trial investigating pharmacodynamic responses to sitagliptin. This sub-study was designed as a crossover study in which healthy individuals underwent a control oral glucose tolerance test (OGTT). After a washout period of at least one week, participants underwent a second OGTT. Two hours prior to the second OGTT, participants received sitagliptin (100 mg, p.o.). On the days of the OGTTs, participants were transported to the Amish Research Clinic, in the fasting state, and their heights, weights, and vital signs were measured. Participants were weighed in a standardized manner while wearing a gown. Reproductive age women underwent a pregnancy test to exclude women who were pregnant. Fasting blood tests were obtained to measure fasting plasma glucose and lipid profile. OGTTs were conducted as described previously ^8^. Two hours after receiving the intervention [either placebo or sitagliptin (100 mg, p.o.)], participants received oral glucose (75 g). Blood samples were obtained at 7 time points: 0, 30, 60, 90, 120, 150, and 180 minutes.

### 2.3 | Eligibility criteria

To be eligible to participate in the clinical trial, individuals were required to be of Amish descent, at least 18 years old, and have BMI between 18-40 kg/m^2^. Exclusion criteria are summarized in the Supplementary Appendix.

### 2.4 | Clinical chemistry

Processing of blood samples and laboratory methods are summarized in the Supplementary Appendix.

### 2.5 | Data and statistical analyses

We established two primary end points: sitagliptin’s effect on area under the curve (AUC) for levels of (a) insulin and (b) glucose during the course of a 3-hour OGTT. We also established several secondary endpoints: sitagliptin’s effect on AUC for intact GLP1, total GIP, and glucagon during the course of the first hour of an OGTT. After having reviewed the OGTT data, we established six exploratory end-points:

- *<u>T30:T60 ratio for insulin</u>*: the ratio of the insulin level at 30 min divided by the insulin level at 60 min in both the (a) control OGTT and (b) sitagliptin OGTT.
- *<u>T30:T60 ratio for glucose</u>*: the ratio of the glucose level at 30 min divided by the glucose level at 60 min in both the (c) control OGTT and (d) sitagliptin OGTT.
- *<u>Drug effects</u>*: (e) the ratio of the T30:T60 ratio for insulin in the sitagliptin OGTT divided by the T30:T60 ratio for insulin in the control OGTT; and (f) the ratio of the T30:T60 ratio for glucose in the sitagliptin OGTT divided by the T30:T60 ratio for glucose in the control OGTT.

Statistical significance was assessed using two-tailed Student’s t-tests for paired data. Although a p-value of p<0.05 was defined as the threshold for nominal statistical significance, a more stringent threshold (e.g., p<0.001) may be appropriate to account for multiple comparisons.

## 3 | Results

### 3.1 | Disposition and adverse events

Disposition of participants and adverse events are summarized in Fig. S1. Fifty-nine individuals were enrolled in this clinical trial between June, 2016 – November, 2018. Forty-seven participants completed oral glucose tolerance tests (OGTT). Research participants’ baseline characteristics are summarized in Table S1.

### 3.2 | Oral glucose tolerance tests: impact of sitagliptin

Sitagliptin decreased the area under the curve (AUC) for glucose by 9% (p=0.0003) but did not significantly affect the AUC for insulin (Fig. 1). Nevertheless, sitagliptin appeared to change the shapes of the curves corresponding to time courses for both glucose and insulin – moving the peaks of the curves for both mean glucose and mean insulin from 60 min (control) to 30 min (post-sitagliptin). Accordingly, we calculated ratios of the levels of glucose or insulin at 30 min divided by the corresponding levels at 60 min (T30:T60) (Fig. 2AB). Sitagliptin increased the mean T30:T60 ratio for glucose from 0.93±0.02 to 1.11±0.04 (p=0.0005) and the mean T30:T60 ratio for insulin from 0.87±0.05 to 1.62±0.04 (p=0.04). We hypothesized that drug-induced increases in early insulin secretion at 30 min drive the subsequent decrease in glucose at 60 min. This hypothesis is supported by the observation that individual values of the T30:T60 ratio for glucose were correlated with individual values of the T30:T60 ratio for insulin (r=0.70, p<0.0001) (Fig. 2C). We observed substantial inter-individual variation in the magnitudes of sitagliptin’s effect on levels of insulin and glucose (Fig. S2). Approximately 30% of participants experienced a numerical decrease in the value of the T30:T60 ratio for insulin after receiving sitagliptin. We hypothesize that these small numerical decreases were likely explained by experimental variation rather than a true “paradoxical” effect of sitagliptin. Similarly, approximately 30% of participants experienced a numerical decrease in the value of the T30:T60 ratio for glucose after receiving sitagliptin. Among the individuals in whom sitagliptin was accompanied by increased T30:T60 ratios, the maximum drug effects were ∼3-fold for glucose and ∼20-fold for insulin.

**FIGURE 1.**
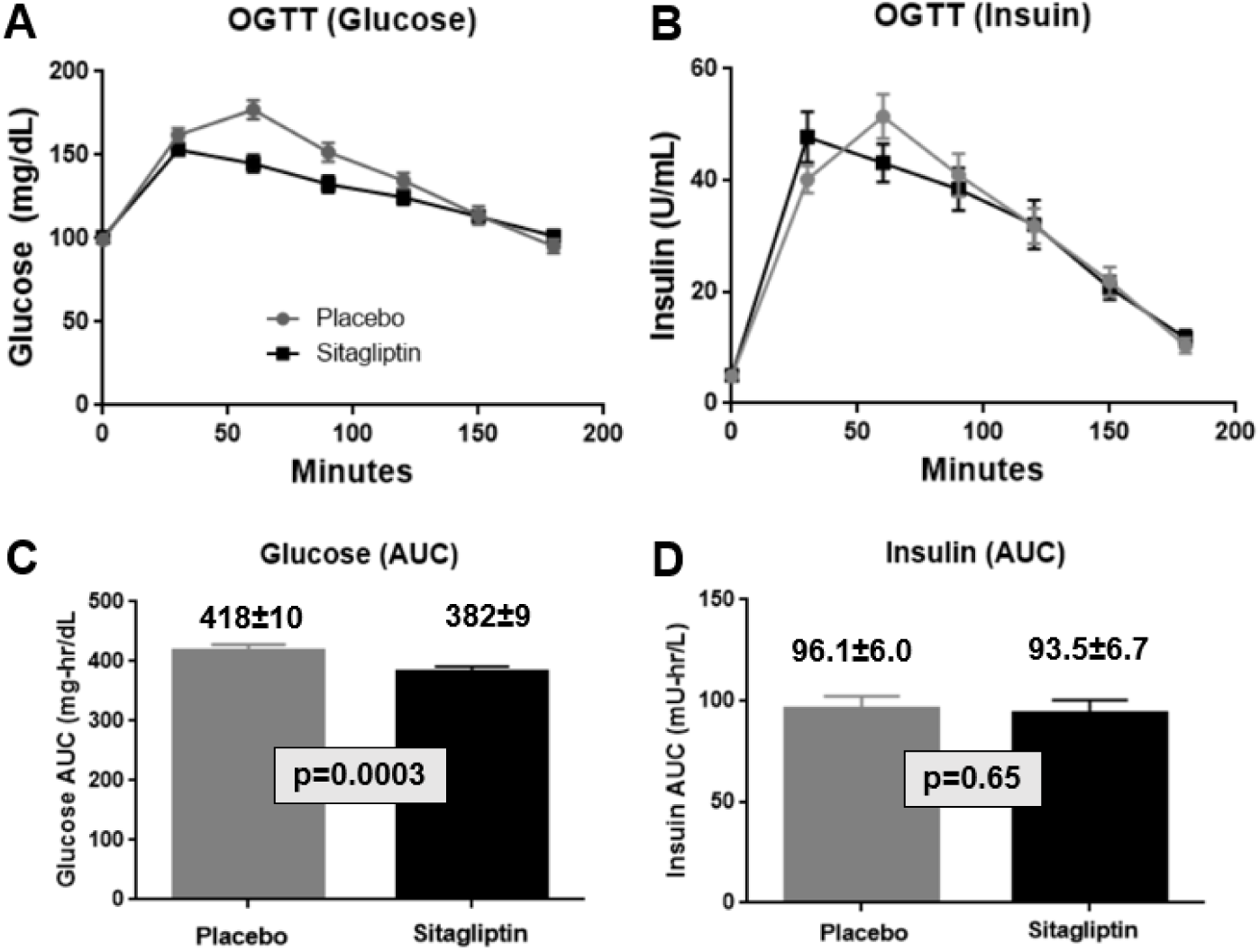
Sitagliptin decreases area under the curve (AUC) for glucose levels during an oral glucose tolerance test (OGTT). Forty-seven participants underwent a control 3-hour OGTT (75 grams of glucose) (depicted by gray circles in panels A and B and gray bars in panels C and D). A second OGTT was conducted one week later. Sitagliptin (100 mg, p.o.) was administered two hours prior to initiating the second 3-hour OGTT (depicted by black squares in panels A and B and black bars in panels C and D). Data on glucose levels are presented in panels A and C. Data on insulin levels are presented in panels B and D. Data are expressed as means ± SEM. p-values (panels C and D) were calculated using a two-tailed Student’s t-test for paired data.

**FIGURE 2.**
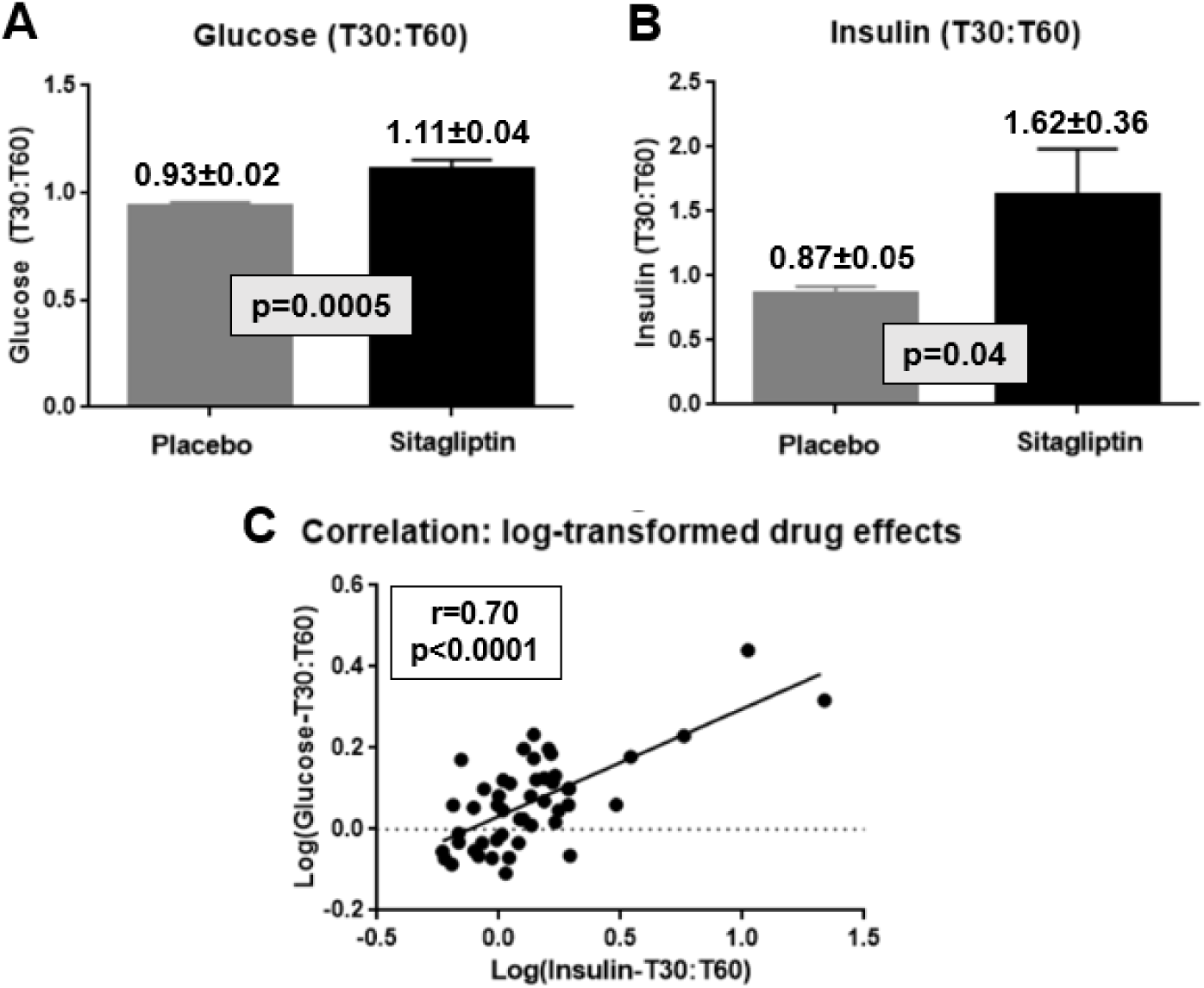
Sitagliptin increases early insulin secretion and improves glucose tolerance during an OGTT. Data from FIG. 1 were reanalyzed by calculating T30:T60 ratios by dividing the concentrations of glucose (panel A) and insulin (panel B) at 30 min by the corresponding concentrations at 60 minutes. Gray bars correspond to data from the control OGTTs (panels A and B); black bars correspond to data from the sitagliptin OGTTs (panels A and B). Data are expressed as means ± SEM. p-values (panels A and B) were calculated using a two-tailed Student’s t-test for paired data. Panel C depicts a graph of individual values of the logarithms of the T30:T60 ratio for glucose as a function of the logarithms of corresponding T30:T60 ratios for insulin.

### 3.3 | Biological variable affecting responses during OGTTs

*<u>Sex</u>*. Mean plasma levels of both glucose and insulin were higher in females than in males during the later time points in OGTTs (90-180 min) (Fig. 3). This sex difference was observed in both control OGTTs and sitagliptin OGTTs. Sitagliptin significantly increased mean T30:T60 ratios for glucose in both males and females (p=0.03). Sitagliptin significantly increased the mean T30:T60 ratio for insulin in males (p=0.01). In addition, the numerical value for the mean T30:T60 ratio for insulin was increased by sitagliptin in females (p=0.11). We did not observe significant differences between the magnitude of the drug effects on mean T30:T60 ratios for either glucose or insulin (Table S2).

**FIGURE 3.**
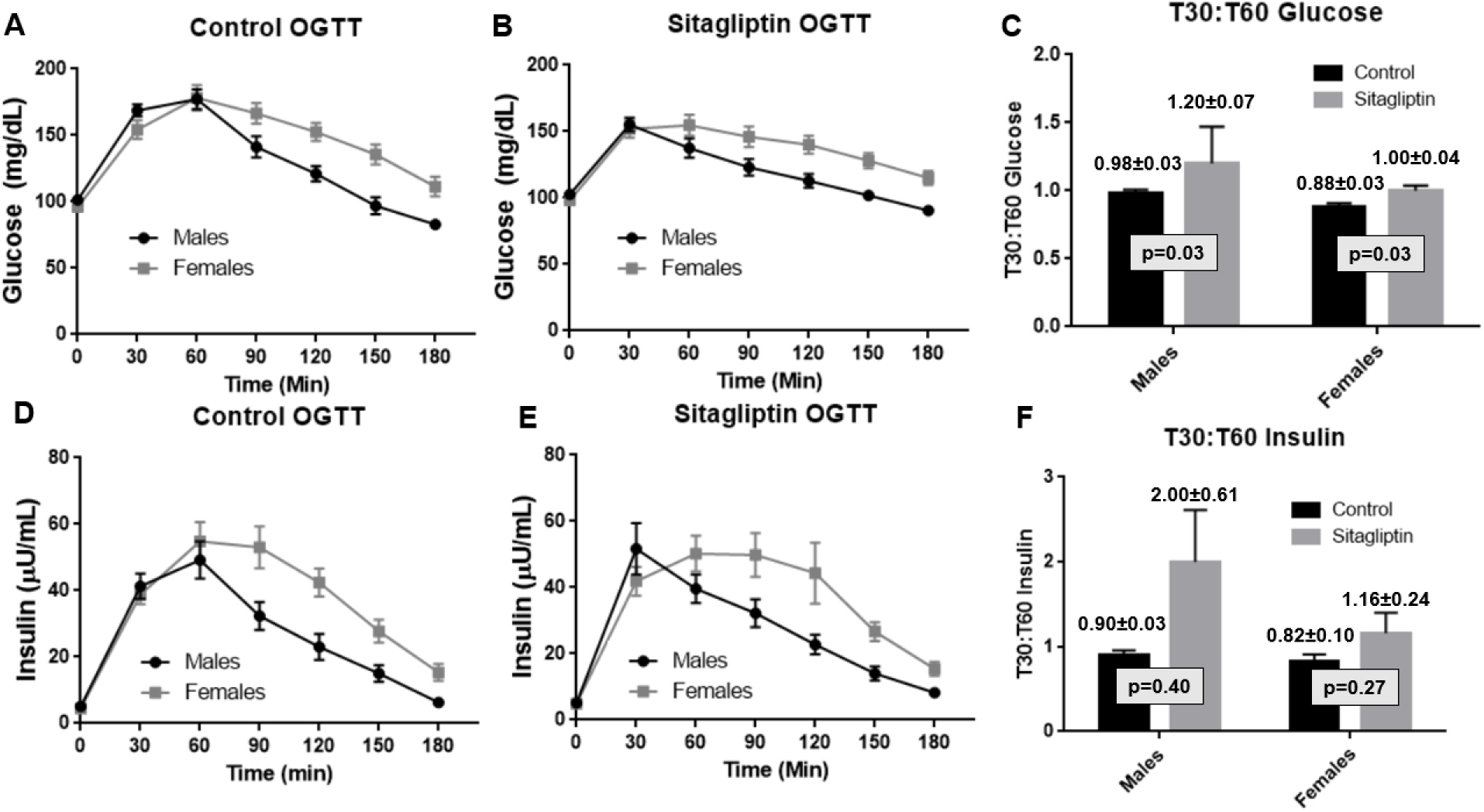
Sex as a biological variable: sex-differences in responses to oral glucose. Data from Figs. 1A and 1B were re-analyzed after stratification according to the self-identified sex of the participant. Data on glucose levels are plotted in Panel A (control OGTT) and Panel B (Sitagliptin OGTT). Data on insulin levels are plotted in Panel D (Control OGTT) and Panel E (Sitagliptin OGTT). Data from Figs. 2A and 2B were re-analyzed after stratification according to the sex of the participant. Data on T30:T60 ratios for glucose and insulin are plotted in Panels C and F, respectively. Data are presented as means ± SEM. p-values were calculated using a two-tailed Student’s t-test for paired data.

*<u>Age</u>*. The variance in age accounted for only 3% of the variation in T30:T60 ratios for glucose in control OGTTs and 14% of the variance in T30:T60 ratios for glucose in sitagliptin OGTTs (Fig. S3). The variance in age accounted for only 4% of the variation in T30:T60 ratios for insulin in control OGTTs and 12% of the variance in T30:T60 ratios for insulin in sitagliptin OGTTs (Fig. S3).

*<u>BMI</u>*. The variance in BMI accounted for only 4% of the variation in T30:T60 ratios for glucose in control OGTTs and 17% of the variance in T30:T60 ratios for glucose in sitagliptin OGTTs (Fig. S4). The variance in BMI accounted for only 0.04% of the variation in T30:T60 ratios for insulin in control OGTTs and 0.5% of the variance in T30:T60 ratios for insulin in sitagliptin OGTTs (Fig. S4).

### 3.4 | Impact of sitagliptin on secretion of GLP1, GIP, and glucagon

Sitagliptin increased mean levels of intact GLP1 at all time points (0, 30, and 60 min) and induced a 2.3-fold increase in the mean AUC for intact GLP1 from 7.0±0.7 to 15.8±1.5 pmol-hr/L (p=3×10^-8^) (Fig. 4A). Sitagliptin increased the mean AUC for glucagon by 66% from 3.74±0.27 to 6.21±0.46 (p=7×10^-9^) (Fig. 4C). The increase in mean AUC for glucagon is primarily a reflection of an increase in mean glucagon levels at time zero. Sitagliptin did not affect mean total GIP levels at time zero but decreased mean AUC for total GIP by 24% (Fig. 4B). We did not conduct assays that specifically measured levels of intact GIP.

**FIGURE 4.**
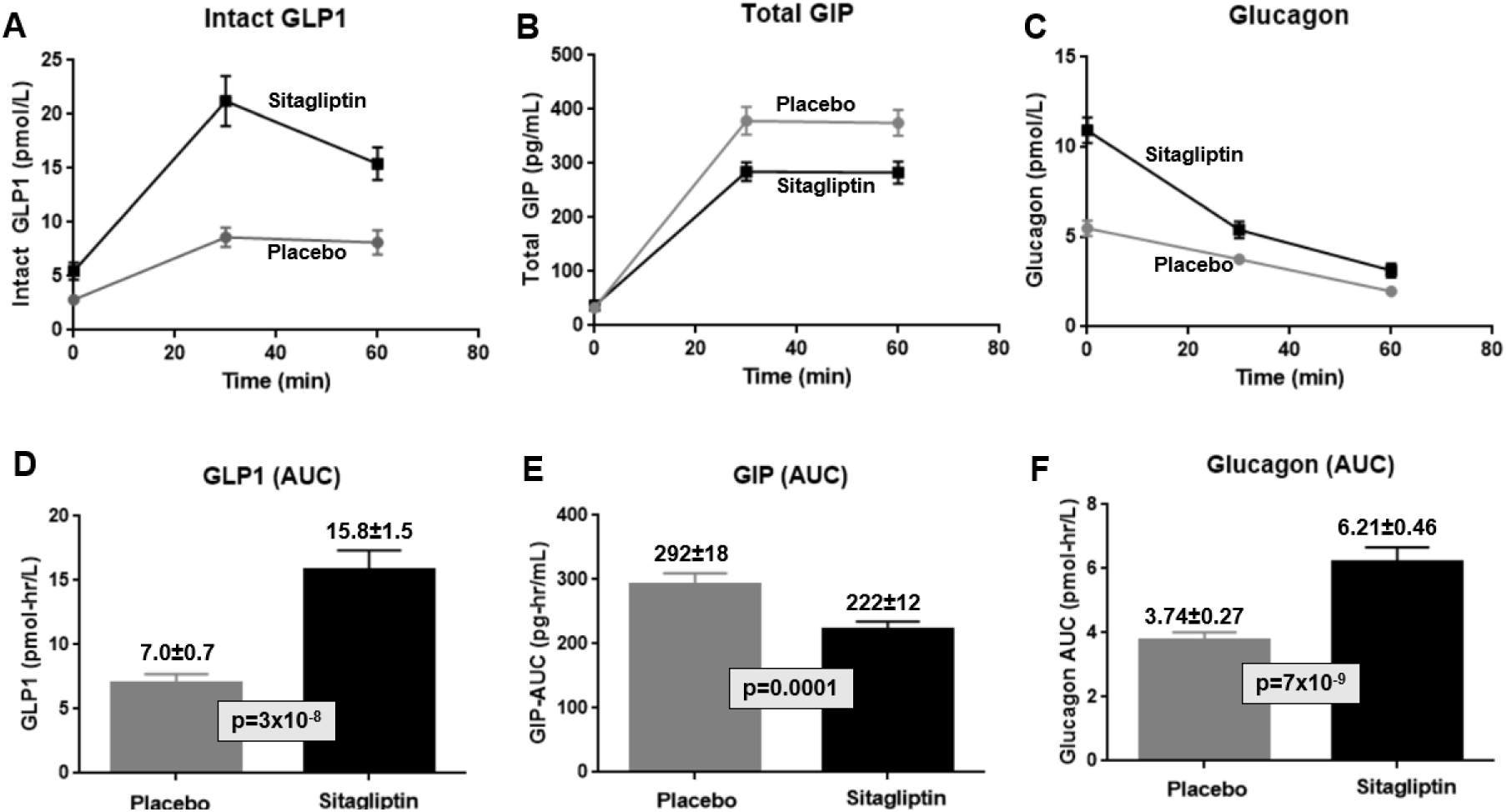
Effects of sitagliptin on areas under the curve (AUC) for levels of intact GLP1, total GIP, and glucagon as a function of time in an oral glucose tolerance test (OGTT). Time courses are depicted for GLP1 (panel A), GIP (panel B), and glucagon (panel C). Areas under the curve are depicted for GLP1 (panel D), GIP (panel E), and glucagon (panel F). Data are expressed as means ± SEM. p-values (panels C and D) were calculated using a two-tailed Student’s t-test for paired data.

We hypothesized that drug-induced increases in GLP1 secretion contributed to the drug-induced increase in insulin secretion during the OGTT. This hypothesis is supported by the observation that individual values of the drug-effect on insulin levels were correlated with individual values of the drug effect on GLP1 levels (r=0.29, p=0.04) (Fig. 5).

**FIGURE 5.**
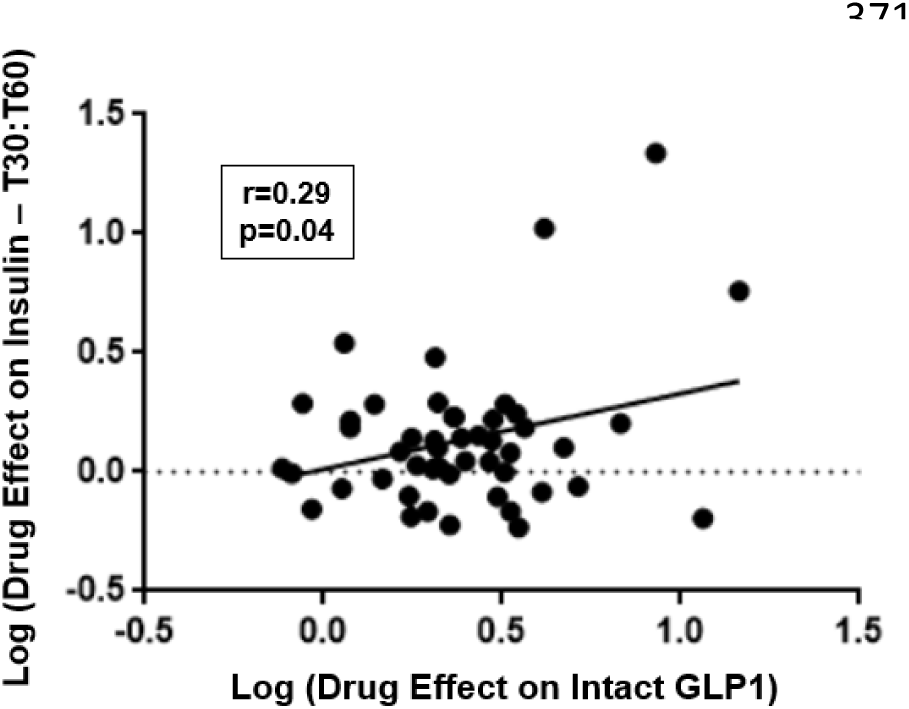
Effects of sitagliptin on insulin secretion are correlated with the magnitude of sitagliptin’s effects on levels of intact GLP1. Individual values of the logarithm of the T30:T60 ratio for insulin are plotted as a function of the logarithms of individual values of the effect of sitagliptin on the AUC for intact GLP1.

As this clinical trial was a sub-study within our pilot clinical trial of pharmacodynamic responses to exenatide ^7^, we inquired whether responses to sitagliptin were correlated with responses to exenatide. Forty-five participants completed both frequently sampled intravenous glucose tolerance tests in our exenatide clinical trial and oral glucose tolerance tests in our sitagliptin clinical trial. The variance in exenatide’s effect to increase first phase insulin secretion accounted for only 0.7% of the variance in sitagliptin’s effect to increase T30:T60 for insulin (r=0.084) (Fig. S5). The variance in exenatide’s effect to accelerate the rate of glucose disappearance accounted for only 0.007% of the variance in sitagliptin’s effect to increase T30:T60 for insulin (r=0.0084) (Fig. S5).

## 4 | Discussion

Our clinical trial demonstrated that sitagliptin (100 mg) decreased mean AUC for glucose by 9% when administered to healthy volunteers two hours prior to initiation of an OGTT (Fig. 1).

Similarly, Herman et al. ^9^ reported that sitagliptin (200 mg) decreased mean weighted averages for *incremental* glucose by 18% when administered to patients with type 2 diabetes two hours before an OGTT. Whereas Herman et al. ^9^ reported that sitagliptin (200 mg) increased mean weighted averages for insulin and C-peptide by ∼20% in patients with type 2 diabetes, our clinical trial did not detect a significant effect of sitagliptin (100 mg) on mean AUC for insulin in healthy volunteers. Both Herman et al. ^9^ and we demonstrated that sitagliptin induced an approximately twofold increase in levels of intact GLP1. Herman et al. ^9^ demonstrated that sitagliptin increased levels of active GIP while inducing a modest decrease in total levels of GIP. Although we did not measure levels of intact GIP, we confirmed their observation that sitagliptin decreased mean AUC for GIP by 24%. As hypothesized by Herman et al., it seems likely that sitagliptin-induced increases in intact GLP1 and/or intact GIP mediate the effect of drug to improve glucose tolerance. Whereas Herman et al. ^9^ reported a 14% decrease in glucagon levels, we observed a highly significant 66% increase in mean AUC for glucagon in response to sitagliptin (100 mg). Although it is unclear how to explain the difference between data in healthy volunteers versus patients with type 2 diabetes, it is noteworthy that diabetes is associated with dysregulation of glucagon secretion ^10–12^. Mathematical modeling of meal tolerance tests demonstrated that vildagliptin increased insulin secretion (primarily in response to breakfast and lunch rather than dinner) without altering glucose sensitivity of the β-cells ^13,14^. Alsalim et al. ^15^ reached similar conclusions when mathematical modeling was applied to analysis of the effect of sitagliptin on meal tolerance tests conducted either in healthy volunteers or patients with type 2 diabetes. Just as we observed that sitagliptin induced a small effect to decrease mean AUC for glucose (p=0.0003) without altering mean AUC for insulin in healthy volunteers, Alsalim et al. similarly observed that sitagliptin decreased mean AUC for glucose by 14% in both healthy volunteers and patients with type 2 diabetes without inducing a statistically significant effect on mean AUCs for insulin or C-peptide.

How can one explain the apparent paradox that sitagliptin improved glucose tolerance (i.e., significantly decreased the mean AUC for glucose) without increasing mean AUC for insulin? Although sitagliptin (100 mg) did not alter mean glucose levels at 0 or 30 min, sitagliptin decreased glucose levels at later time points in the OGTT – most notably at 60 and 90 min. Moreover, although sitagliptin increased mean insulin levels at 30 min, mean insulin levels at 60 min were actually lower for the sitagliptin OGTTs than for the control OGTTs. Mean AUCs were unchanged because the lower levels at 60 min are balanced out by higher levels at 30 min. We hypothesize that the increase in early insulin secretion (i.e., at 30 min) may drive the subsequent decrease in mean glucose levels (e.g., at 60 and 90 min). This could be viewed as the mirror image of the natural history of type 2 diabetes, in which early insulin secretion (first phase) is selectively lost early in the natural history of the disease. In that context, a selective increase in early insulin secretion addresses the pathophysiology of type 2 diabetes. Based on our observations, we hypothesized that ratios of insulin levels at 30 min divided by insulin levels at 60 min would provide a useful index of the shape of the curves for the time course of insulin secretion. Because sitagliptin increased mean insulin levels at 30 min and decreased mean insulin levels at 60 min (Fig. 1B), sitagliptin administration induced a 1.9-fold increase (p=0.04) in the mean T30:T60 ratio for insulin (Fig. 2B). Similarly, because sitagliptin decreased mean glucose levels at 60 min (Fig. 1A), sitagliptin induced a 19% increase (p=0.0005) in the T30:T60 ratio for glucose (Fig. 2A).

The principal objective of our study was to validate pharmacodynamic endpoints for a future pharmacogenomic study of individual responses to a single administration of sitagliptin. Our observations suggest that T30:T60 ratios for glucose and insulin would provide sensitive endpoints for such a pharmacogenomic study. Whereas administration of sitagliptin did not change the AUC for insulin in an OGTT, sitagliptin elicited a substantial (90%) increase in the mean T30:T60 ratio for insulin. Furthermore, we observed substantial inter-individual variation in the magnitude of sitagliptin’s effect on T30:T60 ratios – ranging from little or no effect on either T30:T60 ratio to a 175% increase in the T30:T60 ratio for glucose and a >10-fold increase in the T30:T60 ratio for insulin (Fig. 3). Furthermore, the variance in the sitagliptin-induced increase in the T30:T60 ratio for insulin accounted for ∼50% of the variance in the sitagliptin-induced increase in the T30:T60 ratio for glucose (Fig. 2) – suggesting that T30:T60 ratios provide physiologically relevant indices of insulin secretion and insulin action. We hypothesize that inter-individual variation in insulin sensitivity and/or glucose effectiveness may also contribute to the unexplained variance in the T30:T60 ratio for glucose. Interestingly, we confirmed the previously reported sex-associated difference in responses during an OGTT ^16^, which was attributed to slower absorption of orally administered glucose in females as compared to males. Thus, it will be important to adjust for sex as a covariate when conducting a GWAS searching for genetic variants associated with the response to sitagliptin.

Type 2 diabetes is a progressive disease – with the impairment in beta cell insulin secretion becoming more severe over time ^3,17^. For example, although first line therapy with metformin may provide acceptable metabolic control early in the course of the disease, it is often necessary to add one or more drugs to restore acceptable metabolic control ^1,3,17,18^. Physicians have quite a number of options when selecting a second drug to prescribe for patients who are inadequately controlled on metformin monotherapy – including, DPP4 inhibitors, SGLT2 inhibitors, GLP1R agonists, sulfonylureas, and pioglitazone among others ^1,3,18,19^.

Pharmacogenomics offers potential to predict each individual’s response to specific diabetes drugs – thereby guiding the choice of optimal therapy for each patient. On average, DPP4 inhibitors provide slightly less mean HbA1c-lowering than several other classes of diabetes drugs ^1,3,4^. However, if a physician could predict that an individual patient would experience above average HbA1c-lowering in response to a DPP4 inhibitor, this would increase the attractiveness of the clinical profile for a DPP4 inhibitor for that patient. The impact of a precision therapeutics approach would be magnified if it were possible to apply genetic information to compare the predicted efficacies of several drug classes for each individual patient. This logic provides a strong rationale for conducting pharmacogenomic research in the most important classes of diabetes drugs – including, DPP4 inhibitors, SGLT2 inhibitors, and GLP1R agonists ^1,18,20^. In this context, it is noteworthy that many of our research participants in this clinical trial underwent studies to assess pharmacodynamic responses to both sitagliptin and exenatide. Interestingly, the variance in responses to exenatide accounted for <1% of the variance in responses to sitagliptin (Fig. S2). This observation suggests that some patients might experience above average responses to a DPP4 inhibitor but below average responses to a GLP1R agonists. Although a substantial investment will be required to fully realize the potential of pharmacogenomics and precision therapeutics, such an approach has the potential to transform the approach to designing therapeutic regimens for patients with type 2 diabetes.

## AUTHORS’ CONTRIBUTIONS

*<u>Conception of the clinical trial and PIs for grants</u>*: ALB, SIT

*<u>Acquisition and analysis of data</u>*: ALB, H-RC, MEM, EAS, SIT, HBW, ZSY

*<u>Establishment of Old Order Amish genotype database</u>*: BDM, ARS

*<u>Preparation of first draft of manuscript</u>*: SIT

*<u>Revising and approving final version of manuscript</u>*: all authors

*<u>Accountability for all aspects of work</u>*: ALB and SIT

## Data Availability

Data will be made available to qualified academic researchers subject to terms specified in a Data Transfer Agreement to protect confidential information of participants.

## Supplementary Appendix

### 1 | Exclusion Criteria

- Known allergy to exenatide
- History of diabetes, random glucose >200 mg/dL, or HbA1c ≥ 6.5%
- Significant cardiac, hepatic, pulmonary, or renal disease or other diseases that the investigator judged would make interpretation of the results difficult or increase the risk of participation
- Seizure disorder
- Pregnant by self-report or known pregnancy within 3 months of the start of study
- Currently breast feeding or breast feeding within 3 months of the start of the study
- Estimated glomerular filtration rate <60 mL/min/1.73m^2^
- Hematocrit <35%
- Liver function tests greater than 2 times the upper limit of normal
- Abnormal TSH
- History of pancreatitis or pancreatic cancer. Personal or family history of medullary carcinoma of the thyroid.

### 2 | Oral glucose tolerance tests (OGTTs)

After an overnight fast, participants were transported to the Amish Research Clinic where OGTTs were performed as described previously (1). Briefly, glucose (75 g, p.o.) was administered; blood samples were obtained at the following seven time points: 0, 30, 60, 90, 120, 150, and 180 min. Sitagliptin (100 mg, p.o.) was administered 2 hrs prior to the second OGTT. Plasma levels of glucose and insulin were measured in blood samples for all seven time points. Plasma levels of intact GLP1, total GIP, and glucagon were measured in blood samples for the first three time points (0, 30, and 60 min).

### 3 | Clinical Chemistry

Screening blood samples were obtained by a research nurse during home visits and collected in test tubes as appropriate for each assay: EDTA anticoagulant (purple top tube) for measurement of hematocrit and HbA1c; heparin anticoagulant (green top tube) for measurement of TSH; gray top tubes containing sodium fluoride and potassium oxalate for measurement of fasting plasma glucose; red top tube for collecting serum samples. After placing gray, purple, and green top tubes on ice, blood samples were transported to the clinical laboratory at the Amish Research Clinic (maximum transport time, 2 hours). After centrifugation (3300 rpm for 10 min), plasma/serum was sent on the same day to Quest Diagnostics for assay. Blood samples for OGTTs were collected in EDTA-containing purple top tubes for measurement of plasma insulin and in EDTA/oxalate-containing gray top tubes for measurement of plasma glucose. Fifty µL of aprotinin solution (20,000 KIU/mL) was added per mL blood for glucagon assays. A DPP4 inhibitor (Millipore) was added to plasma samples for assays of GLP1 and GIP at a concentration of 10 µL DPP4 inhibitor per mL blood. Glucose was measured in duplicate using a YSI glucose analyzer. Insulin was assayed in duplicate following the manufacturer’s directions using reagents in kits (#10-1113-01) purchased from Mercodia Inc. The following kits were used for assays of other peptides: intact GLP1 (Millipore #EGLP-35K), total GIP (Millipore #EXHGIP-54K), and glucagon (Mercodia #10-1271-01).

**Table S1.** Demographics and baseline characteristics of study population.

| **Mean ± SEM (range)** | **Males** | **Females** | **Total** |
| --- | --- | --- | --- |
| **N (sample size)** | 27 | 20 | 47 |
| **Age (years)** | 44.3 ± 2.2 | 51.8 ± 2.2 | 47.5 ± 1.6 |
| **BMI (kg/m<sup>2</sup>)** | 27.5 ± 0.7 | 28.7 ± 0.8 | 28.0 ± 0.5 |
| **HbA1c (%)** | 5.49 ± 0.07 | 5.59 ± 0.07 | 5.53 ± 0.05 |
| **Serum creatinine (mg/dL)** | 0.90 ± 0.02 | 0.74 ± 0.02 | 0.83 ± 0.02 |
| **eGFR (mL/min/1.73 m<sup>2</sup>)** | 101.0 ± 2.5 | 94.1 ± 3.6 | 98.0 ± 2.1 |
| **Hematocrit (%)** | 43.8 ± 0.5 | 39.0 ± 0.5 | 41.8 ± 0.5 |
| **Aspartate amino transferase (U/L)** | 18.5 ± 0.9 | 17.4 ± 0.9 | 18.0 ± 0.6 |
| **Alanine amino transferase (U/L)** | 20.7 ± 1.7 | 17.3 ± 1.2 | 19.3 ± 1.0 |
| **TSH (mIU/L)** | 1.82 ± 0.20 | 1.81 ± 0.22 | 1.82 ± 0.14 |

**TABLE S2.** Effect of sitagliptin on T30:T60 ratios for glucose and insulin: data stratified based on self-identified sex. The magnitude of drug effects were calculated by dividing the T30:T60 ratio during the sitagliptin OGTT by the corresponding value for the control OGTT. Data are expressed as means ± SEM for 47 participants (27 males and 20 females). P-values were calculated using two-tailed Student’s t-test for paired data for the T30:T60 ratio for glucose and the log(T30:T60 ratio for insulin).

| Effect of sitagliptin | Males | Females | p-value |
| --- | --- | --- | --- |
| T30:T60 ratio (glucose) | 1.26 ± 0.08 | 1.16 ± 0.06 | 0.36 |
| T30:T60 ratio (insulin) | 2.44 ± 0.82 | 1.46 ± 0.26 | 0.35 |
| N | 27 | 20 |  |

**TABLE S3.** Association of self-identified sex with areas-under-the-curve (AUC) for glucose in oral glucose tolerance test. Areas-under-the-curve for glucose and insulin were calculated using the trapezoidal rule. Data are presented as means ± SEM. p-values were calculated using two-tailed Student’s t-test for either unpaired (association with sex) or paired data (effect of sitagliptin).

**A. Association with sex**
| AUC | Treatment | Males | Females | p-value (for assoc. with sex) |
| --- | --- | --- | --- | --- |
| Glucose | Control | 2393 ± 72 | 2673 ± 94 | 0.02 |
| Glucose | Sitagliptin | 2181 ± 66 | 2465 ± 94 | 0.01 |
| Insulin | Control | 526 ± 45 | 666 ± 52 | 0.06 |
| Insulin | Sitagliptin | 501 ± 48 | 670 ± 72 | 0.05 |
| N |  | 27 | 20 |  |

**B. Effect of sitagliptin**
| AUC | Sex | Control | Sitagliptin | p-value (for effect of sitagliptin) |
| --- | --- | --- | --- | --- |
| Glucose | Male | 2393 ± 72 | 2181 ± 66 | 0.0002 |
| Glucose | Female | 2673 ± 94 | 2465 ± 94 | 8.3x10<sup>-10</sup> |
| Insulin | Male | 526 ± 45 | 501 ± 48 | 0.61 |
| Insulin | Female | 666 ± 52 | 670 ± 72 | 0.95 |
| N |  | 27 | 20 |  |

**Figure S1.**
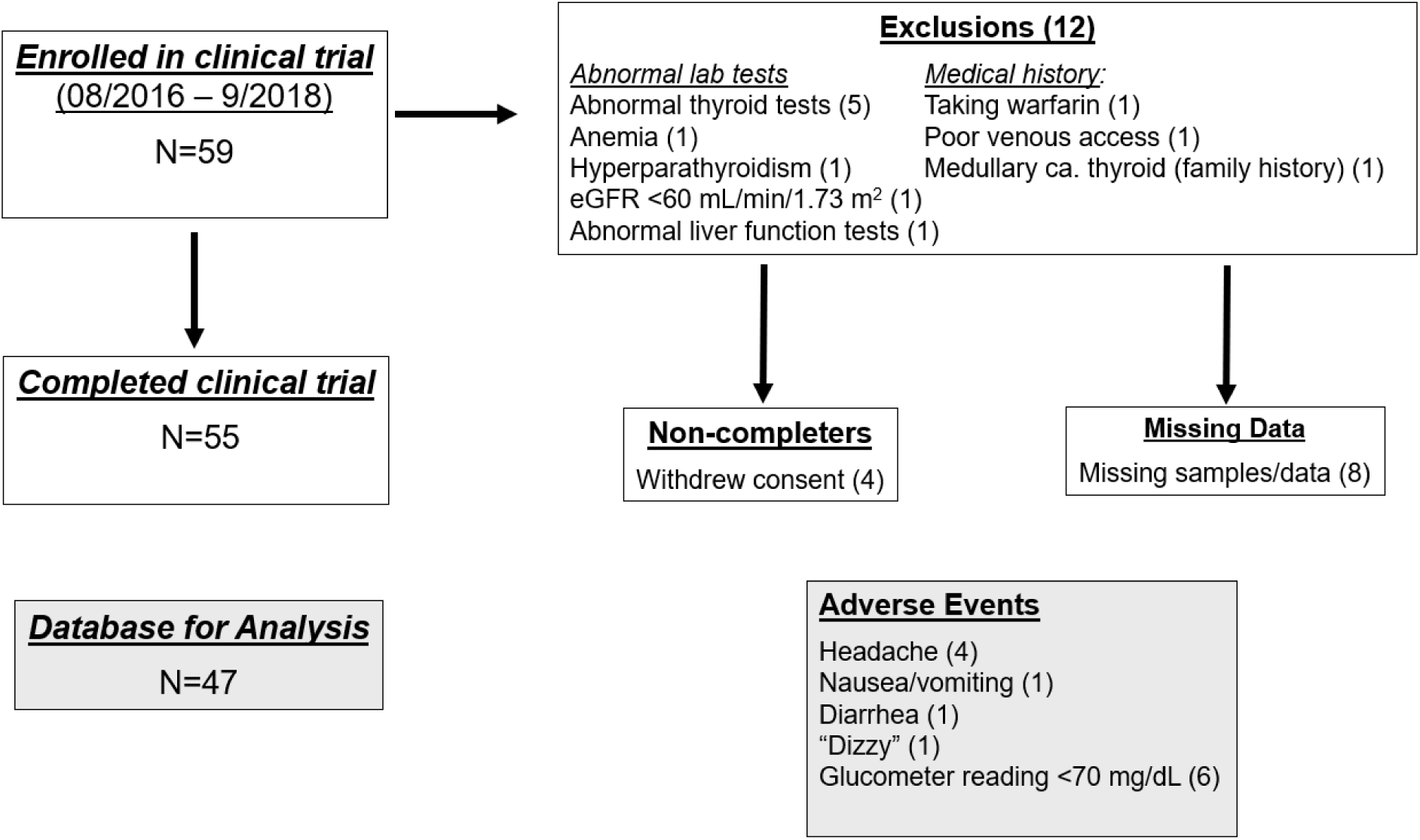
CONSORT diagram summarizing disposition of research participants. Seventy-one individuals underwent screening for the clinical trial between August, 2016 – November, 2018. Twelve enrollees were excluded for the following reasons (Fig. S1): low hematocrit (N=1), abnormal TSH levels (N=5), abnormal liver function tests (N=1), family history of medullary carcinoma of the thyroid (N=1), primary hyperparathyroidism (N=1), low eGFR (N=1), poor venous access (N=1), and treatment with warfarin (N=1). Thus, 59 individuals were judged to be eligible for the clinical trial. Three individuals changed their minds and withdrew from the study after providing informed consent but before undergoing an intravenous glucose tolerance test; technical challenges with establishing venous access led to withdrawal of an additional participant. Fifty-five participants completed two oral glucose tolerance tests. Eight individuals were excluded from the database because of missing data. Thus, data from 47 individuals comprised the final database for the analyses. There were 13 reported adverse events of mild severity. One participant reported feeling light-headed or “dizzy”. There were six events in which a participant’s plasma glucose (determined using a glucometer) was <70 mg/dL during the course of an OGTT. Four participants reported headaches; one reported diarrhea; and one reported nausea.

**FIGURE S2.**
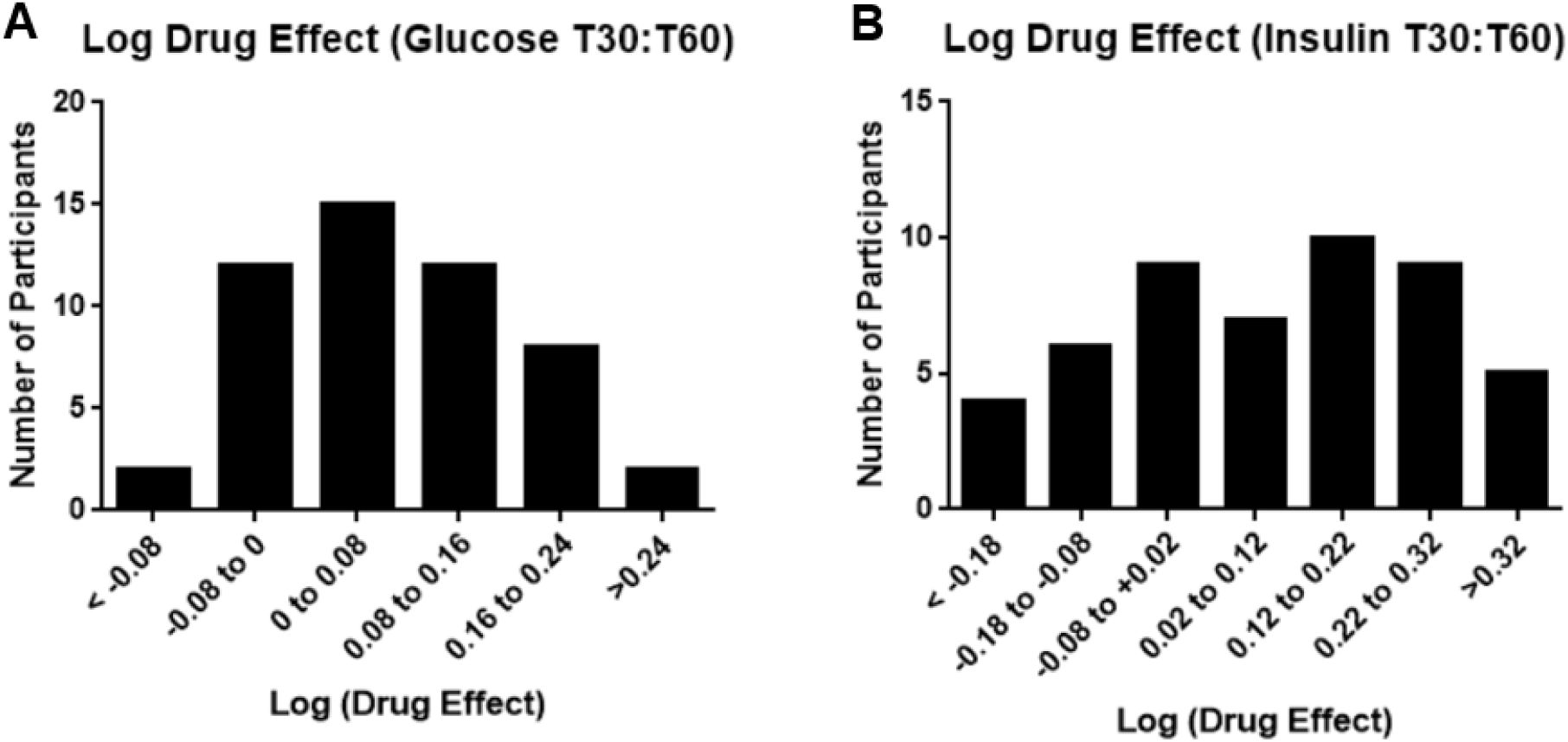
Indices for the effects of sitagliptin were calculated as the ratios of T30:T60 ratio from the sitagliptin OGTT divided by the corresponding ratio from the control OGTT. Analyses were conducted on logarithmically transformed ratios of drug effects for sitagliptin on glucose (panel A) and insulin (panel B). The graphs depict the numbers of individuals whose log(drug effects) fall into the ranges depicted on the respective x-axes.

**FIGURE S3.**
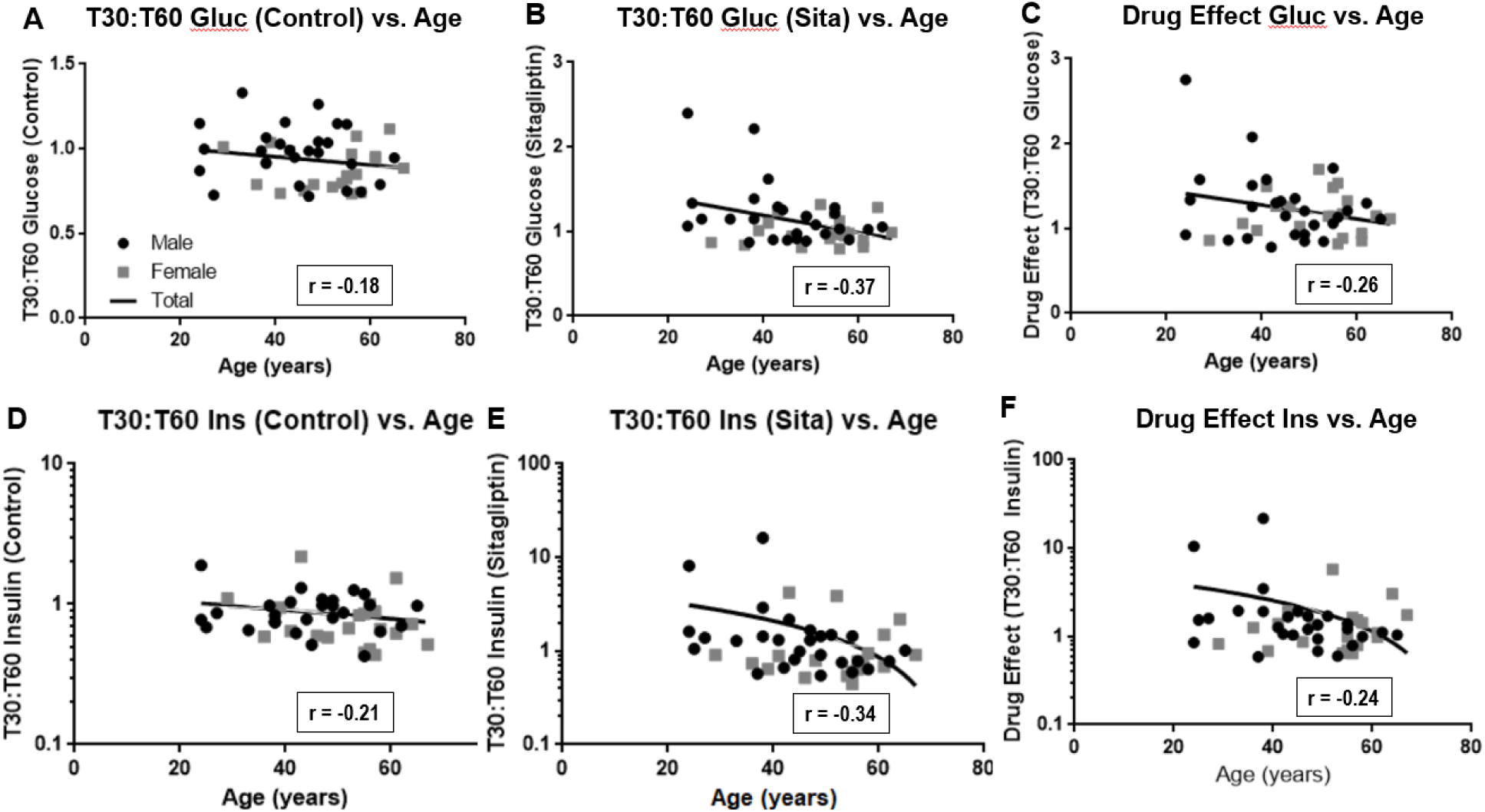
Association of age with responses to sitagliptin. This Figure presents an analysis of associations of age with data on T30:T60 ratios for glucose (panels A-C) and insulin (panels D-F). Data for males and females are depicted by black circles or gray squares, respectively. Data for T30:T60 ratios for glucose are plotted on a linear scale; data for T30:T60 ratios for insulin are plotted on a logarithmic scale. Each panel includes the least squares linear fit to the data. Correlation coefficients (r) were calculated using the data corresponding to T30:T60 ratios for glucose (panels A and B) or logarithms of the T30:T60 ratios for insulin (panels D and E). The drug effect for T30:T60 ratio for glucose was calculated as the ratio of the T30:T60 ratio for glucose in the control OGTT divided by the T30:T60 ratio in the sitagliptin OGTT (panel C). The drug effect for T30:T60 ratio for insulin was calculated as the difference when the T30:T60 ratio for insulin in the sitagliptin OGTT was subtracted from the T30:T60 ratio in the control OGTT (panel F).

**FIGURE S4.**
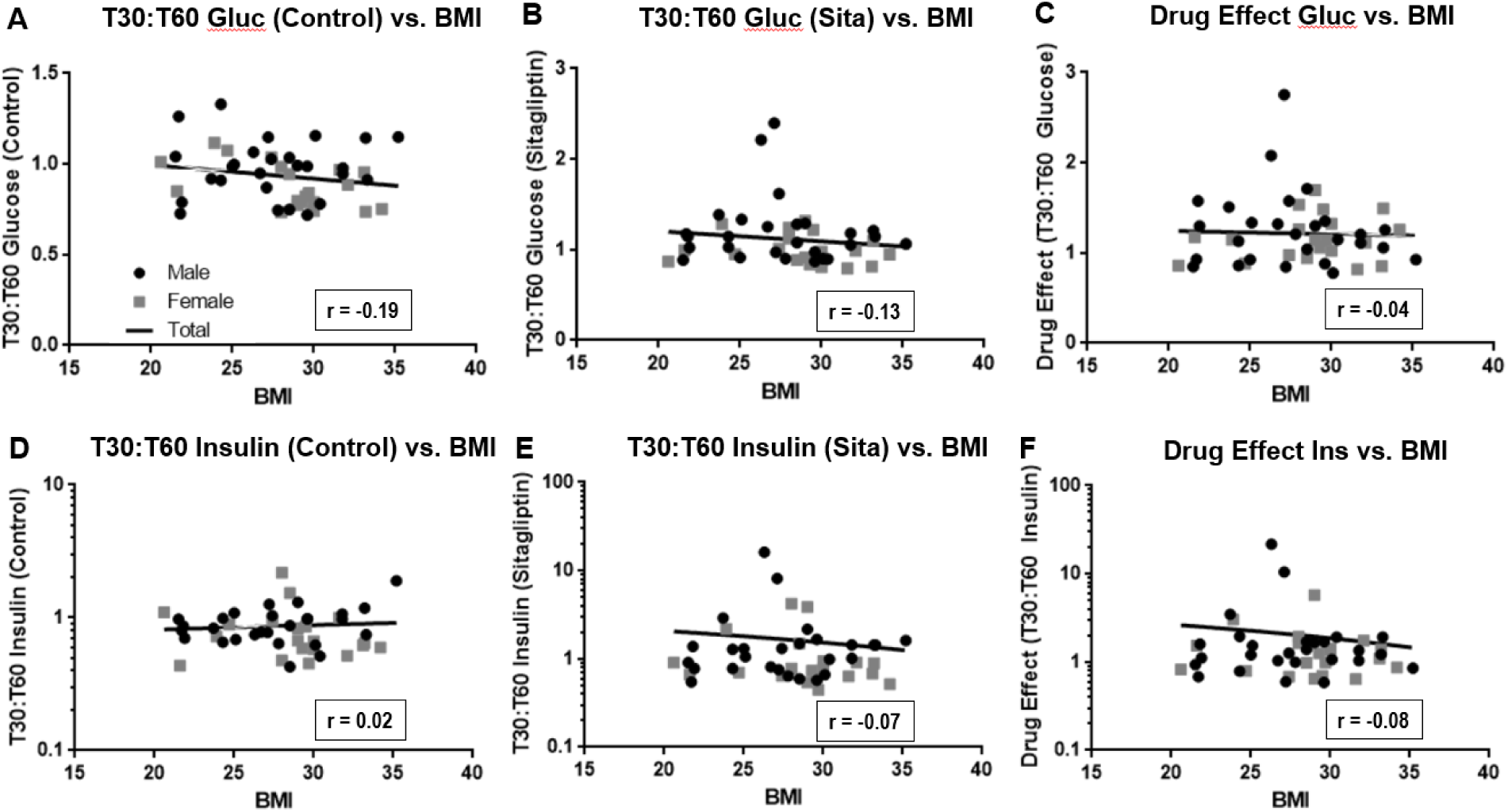
Association of BMI with responses to sitagliptin. This Figure presents an analysis of associations of BMI with data on T30:T60 ratios for glucose (panels A-C) and insulin (panels D-F). Data for males and females are depicted by black circles or gray squares, respectively. Data for T30:T60 ratios for glucose are plotted on a linear scale; data for T30:T60 ratios for insulin are plotted on a logarithmic scale. Each panel includes the least squares linear fit to the data. Correlation coefficients (r) were calculated using the data corresponding to T30:T60 ratios for glucose (panels A and B) or logarithms of the T30:T60 ratios for insulin (panels D and E). The drug effect for T30:T60 ratio for glucose was calculated as the ratio of the T30:T60 ratio for glucose in the control OGTT divided by the T30:T60 ratio in the sitagliptin OGTT (panel C). The drug effect for T30:T60 ratio for insulin was calculated as the difference when the T30:T60 ratio for insulin in the sitagliptin OGTT was subtracted from the T30:T60 ratio in the control OGTT (panel F).

**FIGURE S5.**
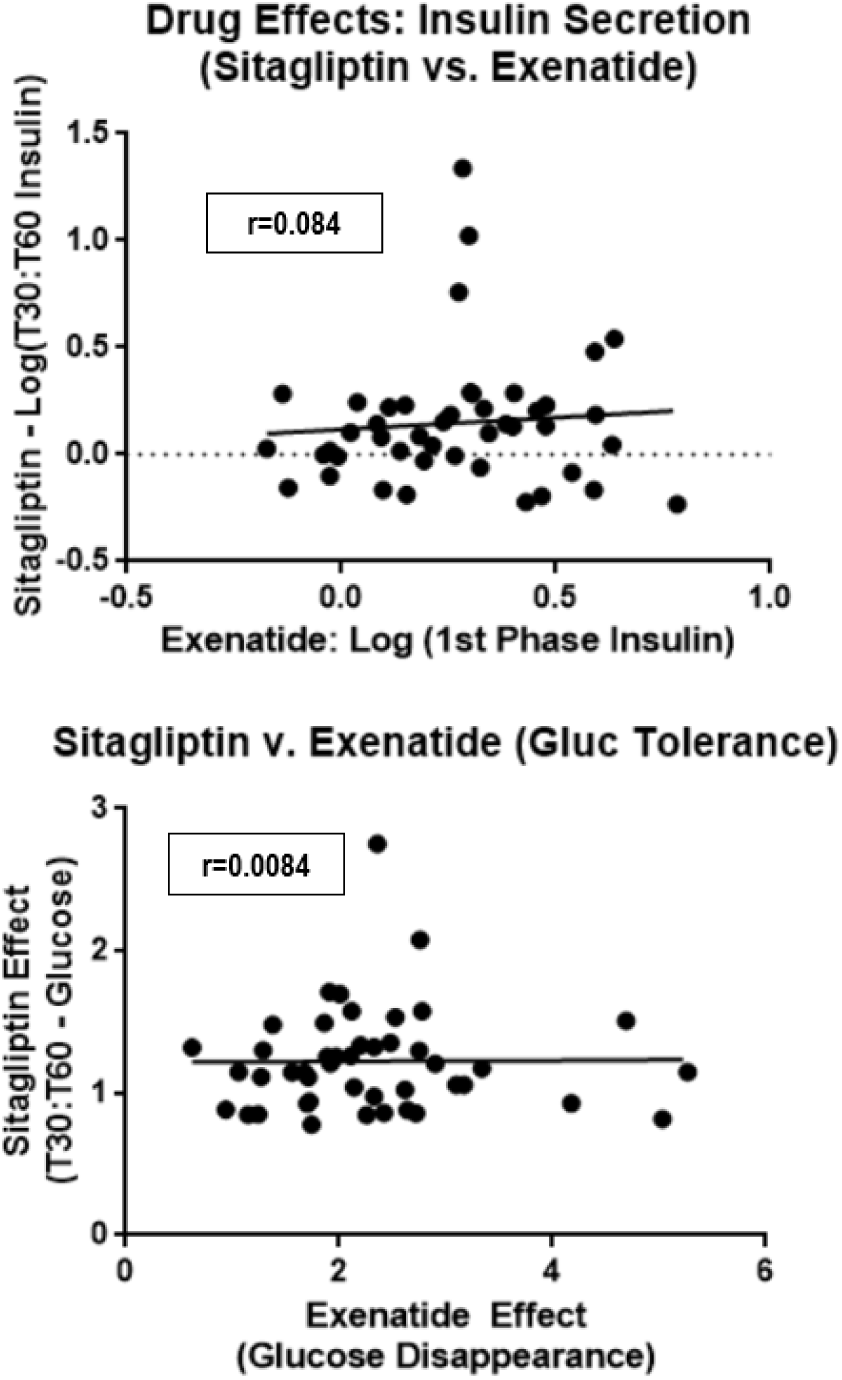
Correlation of pharmacodynamic effects of sitagliptin with pharmacodynamic effects of exenatide. The effect of sitagliptin on log(T30:T60 for insulin) is plotted as a function of the effect of exenatide on log(1^st^ phase insulin secretion) is plotted in the upper panel. The effect of sitagliptin on T30:T60 for glucose is plotted as a function of the effect of exenatide on the glucose disappearance rate is plotted in the lower panel. Data on pharmacodynamic effects of exenatide are based on data from the parent clinical trial with exenatide. Correlation coefficients were calculated using software provided in Microsoft EXCEL.

